# Beyond the Black Box: Avenues for Transparency in Regulating Radiological AI/ML-enabled SaMD via the FDA 510(k) Pathway

**DOI:** 10.1101/2024.07.12.24309602

**Authors:** Alaa Youssef, David Fronk, John Nicholas Grimes, Lina Cheuy, David B. Larson

## Abstract

**Background:** The majority of AI/ML-enabled software as a medical device (SaMD) has been cleared through the FDA 510(k) pathway, but with limited transparency on algorithm development details. Because algorithm quality depends on the quality of the training data and algorithmic input, this study aimed to assess the availability of algorithm development details in the 510(k) summaries of AI/ML-enabled SaMD. Then, clinical and/or technical equivalence between predicate generations was assessed by mapping the predicate lineages of all cleared computer-assisted detection (CAD) devices, to ensure equivalence in diagnostic function.

**Methods:** The FDA’s public database was searched for CAD devices cleared through the 510(k) pathway. Details on algorithmic input, including annotation instructions and definition of ground truth, were extracted from summary statements, product webpages, and relevant publications. These findings were cross-referenced with the American College of Radiology–Data Science Institute AI Central database. Predicate lineages were also manually mapped through product numbers included within the 510(k) summaries.

**Results:** In total, 98 CAD devices had been cleared at the time of this study, with the majority being computer-assisted triage (CADt) devices (67/98). Notably, none of the cleared CAD devices provided image annotation instructions in their summaries, and only one provided access to its training data. Similarly, more than half of the devices did not disclose how the ground truth was defined. Only 13 CAD devices were reported in peer-reviewed publications, and only two were evaluated in prospective studies. Significant deviations in clinical function were seen between cleared devices and their claimed predicate.

**Conclusion:** The lack of imaging annotation instructions and signicant mismatches in clinical function between predicate generations raise concerns about whether substantial equivalence in the 510(k) pathway truly equates to equivalent diagnostic function. Avenues for greater transparency are needed to enable independent evaluations of safety and performance and promote trust in AI/ML-enabled devices.

## Introduction

Artificial intelligence/machine learning (AI/ML) software has advanced rapidly, particularly within radiology. In fact, of the 500+ currently approved AI/ML devices, the vast majority are considered radiological medical devices (1–3). However, despite the wide range of devices available, implementation of these devices into the clinical environment has been slow, in part due to concerns regarding their safety and effectiveness (4,7,8). This lack of trust may be partly because AI/ML devices are often seen as “black boxes” that provide an output with limited transparency on how the device operates or was trained (6,9).

In the United States, the Food and Drug Administration (FDA) considers most AI/ML-enabled software as a medical device (SaMD) to be eligible for the 510(k) premarket submission process as moderate risk (class II) devices. In the 510(k) process, the FDA determines if a device has reasonable assurance of safety and effectiveness by determining if it is substantially equivalent to a legally marketed device (predicate). This process also allows developers to submit their devices with lower levels of evidence, avoiding the longer de novo approval process (10,11). However, the 510(k) process has faced scrutiny for its lack of transparency (10,12,13). Ebrahimian et al. have raised issues regarding the absence of essential information in FDA summaries, while Wu et al. described a concerning lack of comprehensive prospective evaluations among FDA-regulated AI/ML algorithms, with 12/130 algorithms falling short in this aspect (14,15). Previous studies have also demonstrated “predicate creep”, in which broad interpretations of substantial equivalence have resulted in new generations of devices being cleared through claimed equivalence to predicate devices despite iterative design changes, resulting in mismatches in function from the original predicate (16).

The FDA’s 510(k) guidelines recommend providing information about the algorithmic input used to train image analysis-related AI/ML algorithms (13). Nonetheless, it remains unknown what specific information regarding the algorithm development process is included in the 510(k) submissions. This gap is particularly significant given the importance of detailed image annotation instructions in understanding and evaluating the reliability and performance of AI/ML algorithms for their intended clinical tasks (17). In fact, high-quality annotation of reference datasets, which relies on high-quality image annotation instructions, has been shown to be key for validating image analysis algorithms(18).

This study addresses the existing gap in research concerning the availability of algorithm development details, specifically image annotation instructions, in the 510(k) summary document of FDA cleared AI/ML-enabled SaMD that require human annotation in the algorithm development process. Additionally, this study maps the full predicate lineage of diagnostic SaMD, including computer-assisted detection (CAD) devices, to assess substantial equivalence.

## Methods

This study included three stages: 1) identifying radiological AI/ML-enabled SaMD cleared through the 510(k) pathway and their characteristics, 2) evaluating the image annotation instructions included in each 510(k) summary, and 3) mapping their predicate lineages (**Figure 1**).

**Figure 1.**
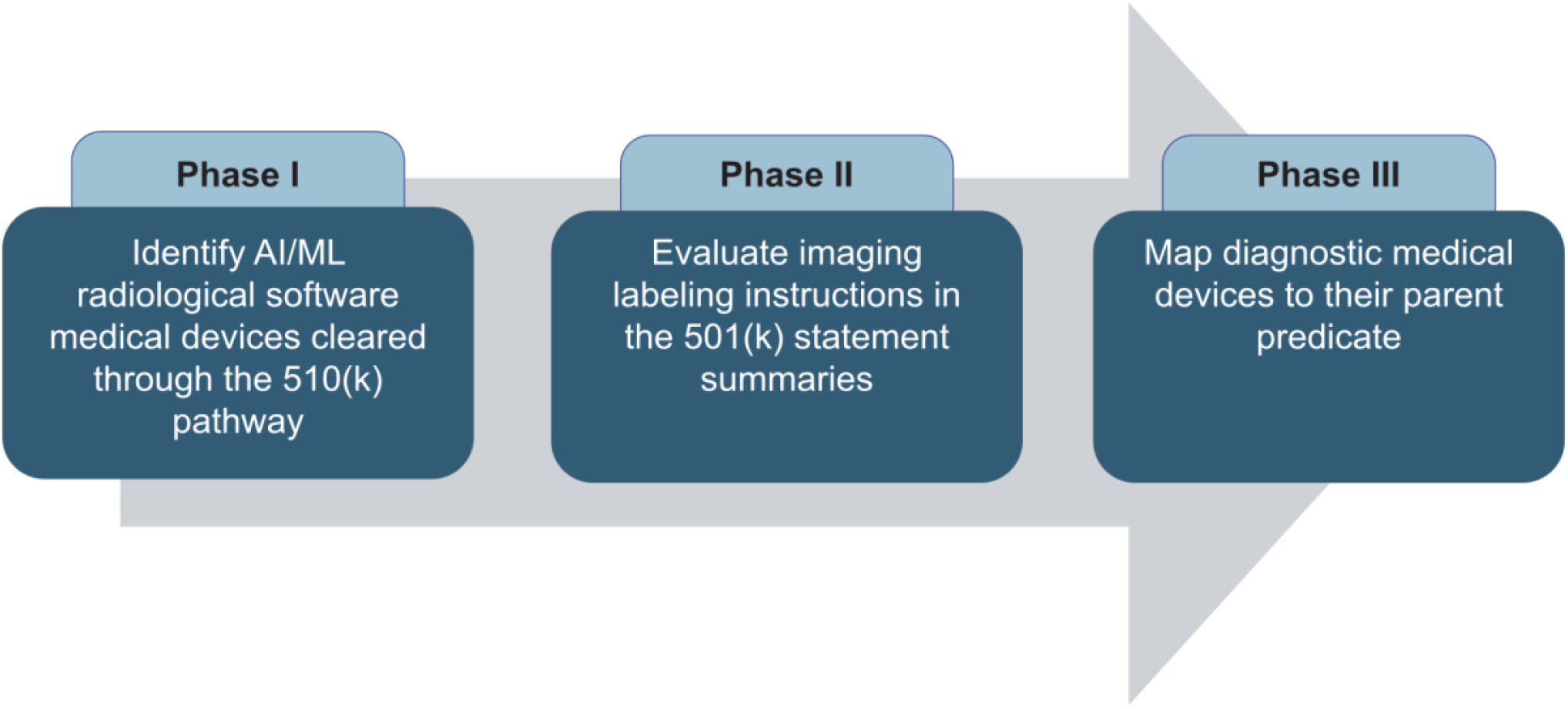
Study methodology for identifying and evaluating diagnositc radiological AI/ML software as a mmedical Device (SaMD) cleared through the FDA 510(k) pathway.

### Identification and Characterization of Radiological AI/ML-based SaMD

The FDA’s public 510(k) database of AI/ML-enabled medical devices was searched from May 15, 2018 to October 30, 2023 to identify radiological AI/ML-enabled SaMD. This study focused only on computer-assisted devices. FDA product codes were used to identify specific computer-assisted devices subtypes: QAS and QFM for CADt (computer-assisted triage) devices, MYN for CADe (computer-assisted detection) devices; POK for CADx (computer-assisted diagnostic) devices; and QDQ and QBS for CADe/x (computer-assisted detection and diagnostic devices (**Table 1**) (3).Although the FDA considers medical imaging management and processing systems (MIMPS) a type of radiological AI/ML-enabled SaMD, MIMPS were excluded from this study because these devices are not all adaptive algorithms and do not all require human-annotated data for algorithm development.

**Table 1.**
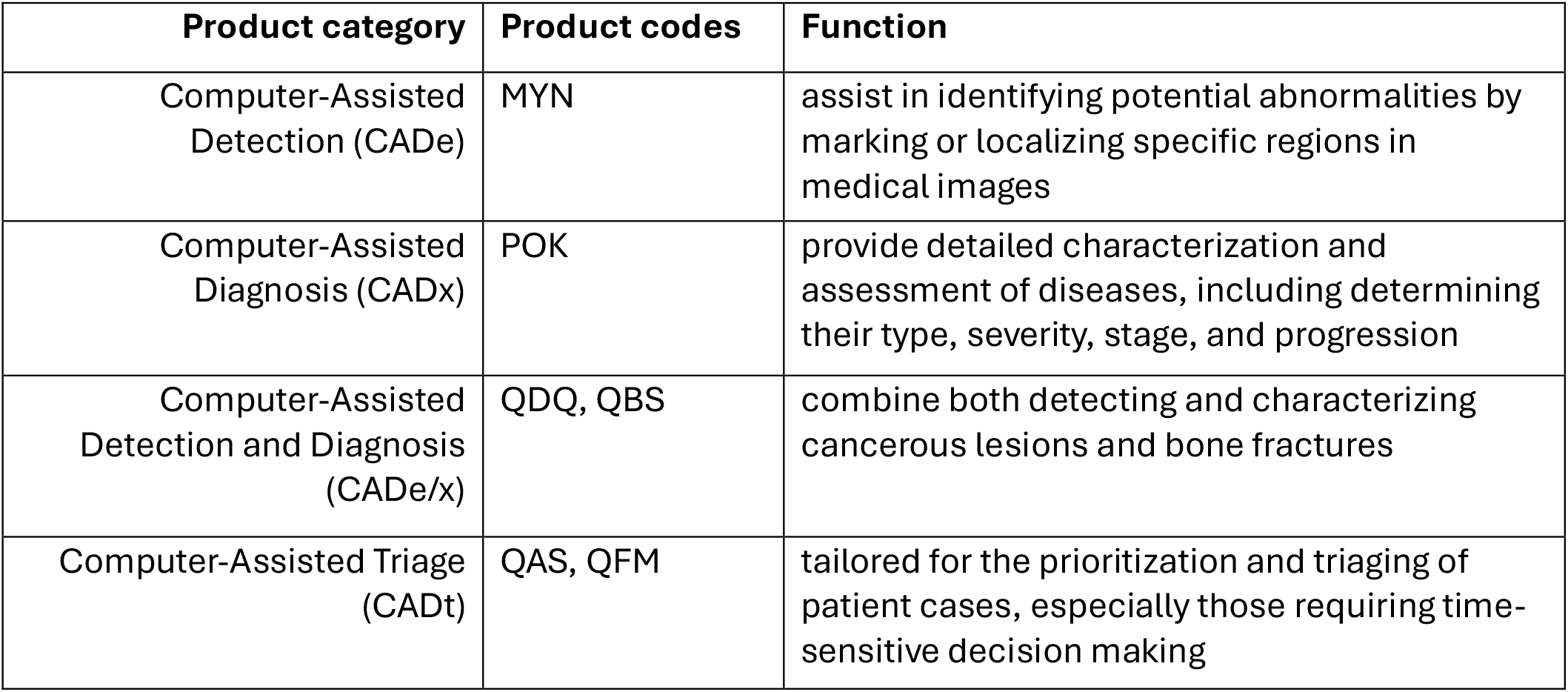
FDA product categories and product codes for Computer-Assisted Devices (CADs)

The following details were extracted from the FDA 510(k) summaries of each cleared device: clearance date, product name, developer, algorithm type, diagnostic function, imaging modality, and algorithmic input details (image annotation instructions or methods). Next, the data were cross-referenced with the AI Central database from the American College of Radiology-Data Science Institute, which is updated more frequently than the FDA database and contains complementary information (19). From AI Central, detailed information about each CAD device, including the applicable medical imaging technique (modality), anatomy, and intended diagnostic function were collected. All data were compiled in Microsoft Excel for analysis.

### Evaluation of Image Annotation Instructions in the 510(k) Summaries

The FDA recommends that all 510(k) summaries include a detailed description of the submitted AI/ML algorithm, including the algorithm input. For this study on medical imaging-focused devices, algorithmic input was defined as image annotation instructions, or the training set of medical images annotated according to definitions provided by experts. These details were considered important for objective evaluation since inconsistency in medical imaging annotations can lead to labeling bias, impacting the accuracy of AI/ML algorithms. To assess the quality of the image annotations instructions provided, a comprehensive review of each device’s 510(k) summary was conducted for guidelines or instructions used to create the annotated images.

Additionally, the 510(k) summaries were reviewed to assess how developers determined the labels’ ‘ground truth’ – a benchmark for measuring an algorithm’s performance. Due to the frequent paucity of details in the 510(k) summaries, product webpages and any peer-reviewed publications listed on the webpages were also searched for data-sharing statements, image annotation instructions, and descriptions of the ground truth.

### Mapping Diagnostic Medical Devices to their Parent Predicate

Predicate lineages were mapped to better understand the similarities between cleared CAD devices and their claimed predicate device. The lineages of all identified CAD devices were manually traced back to their parent predicate by identifying the submission number (K number) of each newly cleared device in the corresponding 510(k) summary until reaching the parent de novo predicate. Each device’s 510(k) summary was manually reviewed to determine its intended purpose, defined as the clinical task the device was developed to perform. All data were extracted into Microsoft Excel, and the resulting map was generated using Adobe Illustrator 2024 version 28.1.

## Results

### Summary of cleared Radiological AI/ML-based SaMD

As of October 31, 2023, the FDA 510(k) Al/ML-enabled software medical devices database listed 298 cleared radiological devices. Of these, 200 (67.1%) were MIMPS, and 98 (32.9%) were CAD devices.

Overall, the number of cleared AI/ML-enabled medical devices grew dramatically from 2008 to 2023 (**Figure 2A**), with MIMPS accounting for the majority of devices cleared through the 510(k) pathway each year. Although the first MIMPS was cleared in 2008, the first CAD device was not cleared through the 510(k) pathway until 2016, with only 4 CAD devices total being cleared between 2016 and 2018. A significant turning point was in 2019, with a substantial increase from 2 CAD devices cleared in 2018 to 10 CAD devices cleared in 2019 alone. This trend continued into 2020, when 18 CAD devices were cleared, and the total number of CAD devices cleared has remained relatively stable from 2020-2023 (16-19 devices cleared/year). Notably, the number of CADt devices cleared/year has steadily grown since 2018, while the proportion of CADe and CADx devices has remained relatively consistent.

**Figure 2.**
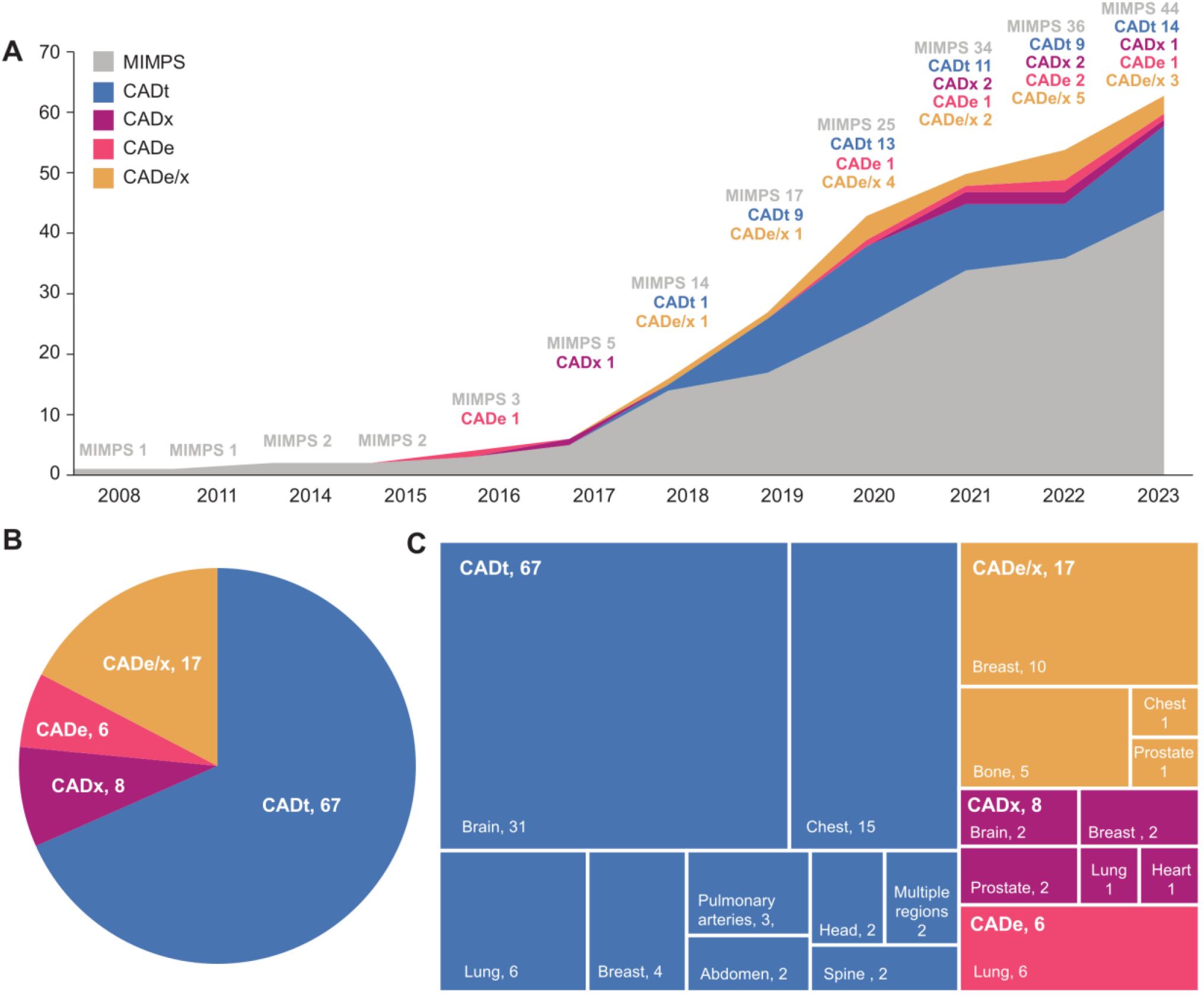
A) Number of cleared radiological AI/ML medical devices by year, showing that medical imaging management processing software ( MIMPS) account for the majority of radiological AI/ML devices cleared through the 510(k) pathway and are driving the increase in AI/ML device clearance rate; B) distribution of the total number of computer-assisted devices (CADs) cleared through the 510(k) pathway from 2008 to 2023, showing most of the cleared CADs were computer-assisted triage (CADt) devices; and C) the distribution of anatomical function among the cleared CADs, illustrating that the cleared CADt, computer-assisted diagnosis (CADx), and computer-assisted detection and diagnosis(CADe/x) devices vary in target anatomy, while all computer-assisted detection (CADe) CADe devices focused on the lungs.

Within the CAD devices, 67 (22.48%) were CADt devices, 6 (2.0%) were CADe devices, 8 (2.68%) were CADx devices, and 17 (5.70%) were CADe/x devices (**Figure 2B**).The intended tasks of the CAD devices varied in anatomical region. The majority of CADt devices (31/67, 46.3%) were designed to triage brain imaging findings, including critical conditions such as large vessel occlusion, intracranial hemorrhage, and subdural hemorrhage. Most CADe/x (10/17, 58.8%) devices focused on the breast, and 2/8 (25%) CADx devices were for breast cancer detection and diagnosis tasks. All 6 CADe devices had applications in the chest region, with the primary function being detecting pulmonary nodules (**Figure 2C**).

### Evaluation of Image Annotation Descriptions in the 510(k) Summaries

None of the 98 devices included image annotation instructions in their 510(k) summaries, on their product web pages, or in any associated publications. Notably, only one device provided access to its training data.

The ground-truth descriptions in the 510(k) summaries also varied. Most devices (56/98, 57.0%) did not provide any information about how the ground truth was defined, while 27 (27.8%) included statements that referenced expert determination, but without details on the experts’ qualifications (**Figure 3**). The objective, quantifiable gold-standard approaches used to determine the ground truth included biopsy (7/98, 7.1%), specific medical device measurements (3/98, 3.2%) and expert application of established clinical standards (i.e., BI-RADS; 5/98, 4.8%). Peer-reviewed publications were found for only 13/98 (13.2%) of the devices (4 CADt, 1 CADe, 1 CADx, and 7 CADe/x). Among these, only two studies were prospective multi-center randomized controlled trials; the rest were retrospective reader studies intended for algorithm validation. None of the publications revealed detailed descriptions or references regarding how the images were annotated to train the algorithm. Only three devices had publications that offered some insights into their annotation definitions. A retrospective multi-reader study evaluating the accuracy of a CADe/x device for breast cancer detection referenced the University of South Florida Digital Database for Mammography Screening, which details breast lesion scoring criteria and links to a publicly accessible ground-truth mammography database for algorithm development and benchmarking (20). In another prospective study for a CADt device for detecting large vessel occlusion, surgical anatomic definitions were applied to CT angiography scans to specify the exact occlusion site (21). Finally, in a retrospective study validating a CADe device for pulmonary nodule detection, the authors defined “solid nodules” based on a published set of standard terms. Of all 13 cleared CAD devices, only these three devices had published studies with clearly defined ground-truths based on acceptable gold-standard approaches.

**Figure 3.**
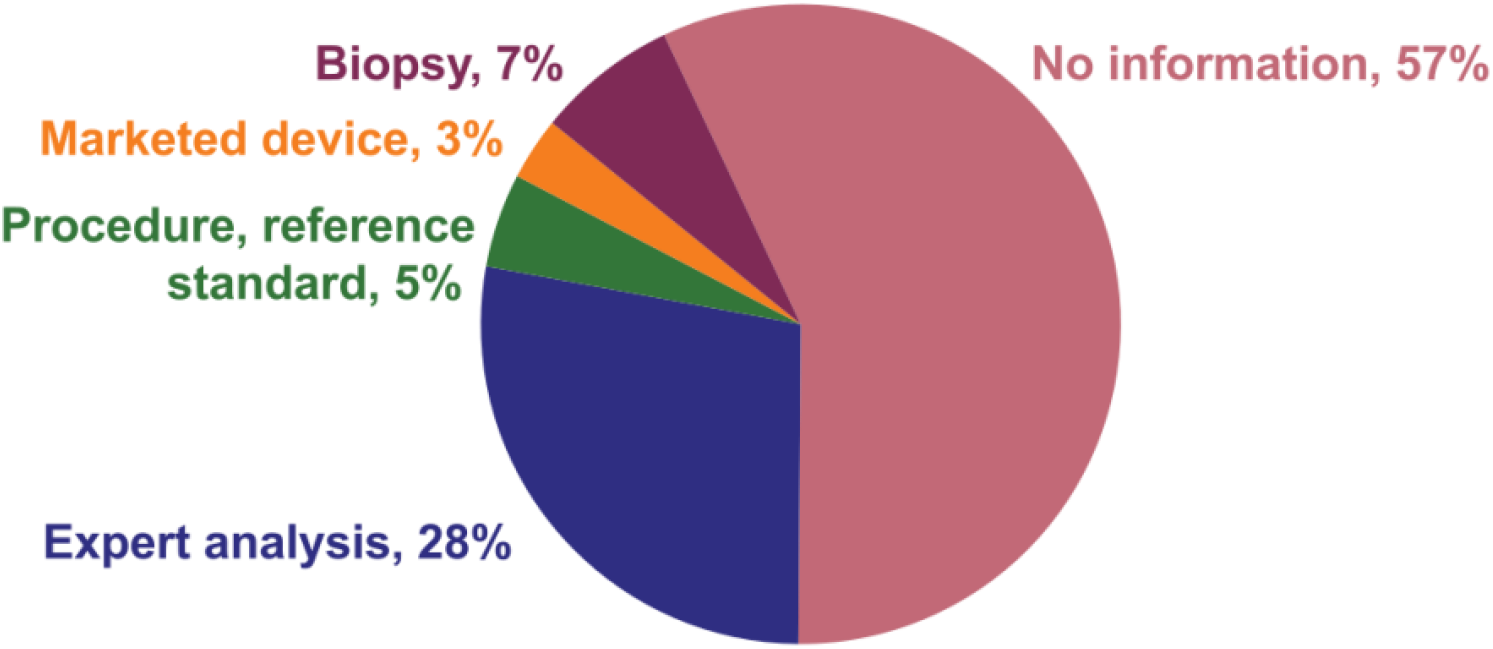
More than half of the 510(k) summaries of all cleared CADs contained no information on how the ground truth used to validate algorithm performance was obtained.

### Mapping Diagnostic Medical Devices to their Parent Predicate

The predicate device lineages of CADt, CADe, CADx, and CADe/x devices cleared through the 510(k) pathway were mapped to ensure similarities in stated diagnostic function. All CADt devices originated from the de novo device Viz.AI (DEN170073), and all CADe devices could be traced back to original predicate RapidScreenTM RS-2000 (P000041), with no mismatches in diagnostic function or imaging modality. However, although all 25 cleared CADx and CADe/x devices originated from two parent predicate devices, QuantX (DEN170022) and OsteoDetect (DEN180005), respectively, there were significant deviations from the claimed predicate.

The first de novo CADx device, QuantX (DEN170022), was cleared in 2017 specifically targeting the detection of breast lesions on mammography. In total, six other CADx devices and two CADe/x devices can be traced back to the same de novo device (**Figure 4A**). Notably, although the initial application focused on mammography, the regulatory scope of the de novo CADx device broadly encompasses any lesions suspicious of cancer as well as other imaging modalities (e.g., US, X-ray, MRI). Following this broad de novo definition, two out of the four first predicate generation devices have diagnostic functions focusing on non-breast lesions with other modalities. For example, the stated function of device Avenda Health AI Prostate Cancer Planning Software (K221624) is to detect prostate lesions on MRI, while that of device OptellumTM Software (K202300) is to detect lung nodules on CT. For another first predicate generation device, Brainomix 360 e-ASPECTS (K200760), the stated function is to detect intracranial hemorrhage on CT, revealing a potential mismatch in the regulatory definition of “lesion suspicious of cancer”. Similar deviations in the de novo regulatory definitions are observed in the subsequent predicate generations. For example, a second-generation device, EchoGo Pro (K201555), for coronary artery disease detection on ultrasound, claimed equivalency to the device Koios DS for Breast (K190442), which also uses ultrasound but aims to detect breast lesions.

**Figure 4.**
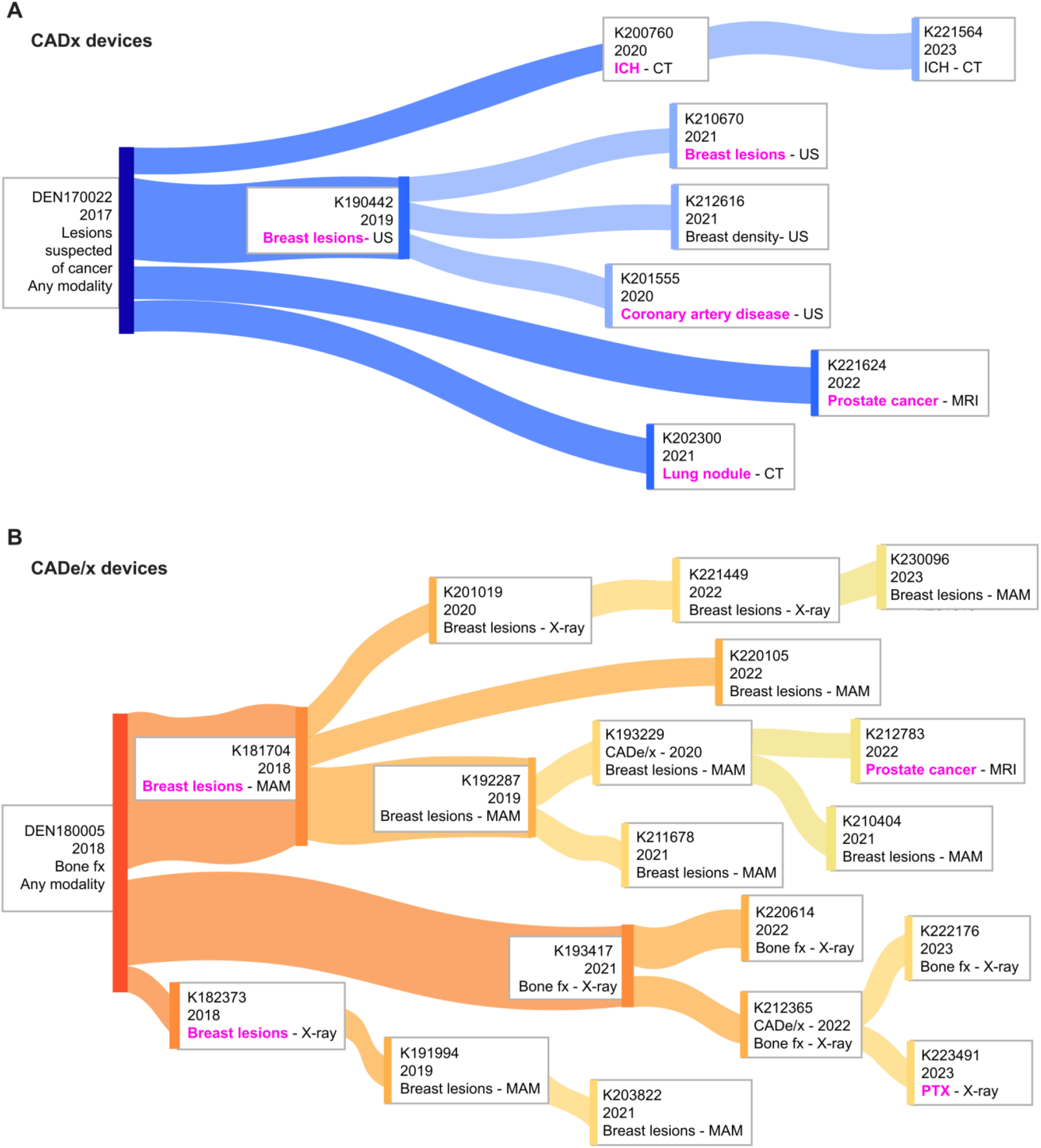
Predicate networks of all A) Computer-Assisted Diagnosis (CADx) (blue tree) and B) Computer-Assisted Detection and Diagnosis (CADe/x)(orange tree) devices approved through the FDA 510(k) pathway, with lighter colors indicating later predicate generations. Pink text denotes where a mismatch in anatomic function and/or imaging modality is present with the cleared device’s claimed predicate device. MAM = mammography, MRI = magnetic resonance imaging, X-ray = radiographic imaging, US = ultrasound, CT = computed tomography, fx = fracture ICH = Intracranial hemorrage, PTX = pneumothorax

In 2018, OsteoDetect (DEN180005) became the first de novo CADe/x device, serving as the parent predicate for all subsequent CADe/x devices cleared through the 510(k) pathway. The FDA classified OsteoDetect (DEN180005) as a device for detecting and diagnosing suspected fracture with any imaging modality (e.g., X-ray, CT, MRI). This de novo DEN180005 served as the direct parent predicate for three first generation devices: FractureDetect (FX) (K193417),Transpara (K181704), and PowerLook Tomo Detection V2 Software (K182373).

However, two of these focus on diagnosing breast cancer lesions (K181704 and K182373), revealed a mismatch in diagnostic function from the defined regulatory scope of suspected fractures (**Figure 4B**). Despite this mismatch to the parent predicate in diagnostic function, Transpara (K181704) for detecting breast lesions on mammography emerged as the most commonly claimed predicate device for later generations of CADe/x devices targeting breast lesion detection, leading to eight breast lesion detection devices total and one prostate cancer detection device. In contrast, only four CADe/x devices— Rayvolve (K220614), FractureDetect (FX) (K193417), BoneView (K212365), and BoneView1.1 US (K222176) — have functions truly equivalent functions to detecting bone fractures similar to the de novo CADe/x device. Later, an additional device, Critical Care(K223491), for detecting pneumothorax, claimed equivalence to the bone fracture detection device BoneVew (K212365) (**Figure 4B**).

These predicate maps reveal that all 25 CADx and CADe/x devices cleared through the FDA 510(k) pathway originate from two parent devices: QuantX (DEN170022), a CADx device for breast lesion detection, and OsteoDetect (DEN180005), a CADe/x for bone fracture detection. Notably, 17 of these 25 devices are intended to detect or diagnose breast lesions, and approximately half (8/17) of these have claimed equivalence to the predicate device Transpara (K181704), which was also designed to detect breast lesions. However, although this breast lesion detection device was cleared through claimed equivalence to the de novo CADe/x , there is a noticeable difference in clinical function between the two devices (bone fracture detection vs. breast lesion detection). These findings demonstrates that claimed equivalence may adhere to the FDA’s current definition of substantial equivalence but is far from true equivalence in diagnostic function.

## Discussion

The rapid development of AI/ML-enabled tools has led to a significant increase in the number of such devices cleared through the FDA 510(k) pathway, with 19 CAD devices cleared in 2023 alone. Notably, although the majority of AI/ML medical devices on the market have been cleared through the 510(k) pathway, this process was not developed specifically for AI/ML-enabled devices, leading to broad interpretations of the required documentation and previous reports of predicate creep in 510(k) AI/ML medical devices (13,16,22). In this study, the 510(k) documents of cleared CAD devices were examined to assess the availability of detailed information regarding the algorithm development process, specifically the image annotation instructions. Additionally, the predicate lineages of AI/ML-enabled SaMD categorized as CAD devices were mapped to evaluate the compatibility of the new devices with their predicates.

There were no descriptions of the annotation instructions used for algorithm development in the 510(k) summaries of the cleared CAD devices or on their product webpages. Over half of the cleared devices also lacked details on how the ground truth was defined for device validation. The importance of clearly defined and validated image annotation instructions to algorithm performance has been widely recognized, as having a clearly defined and known ground truth enables users to objectively assess performance metrics such as algorithm accuracy, reproducibility, and reliability (14,15,18,23,24) Although the FDA is not required to publicly disclose all information, since manufacturers almost never publish this elsewhere, this lack of transparency of the algorithmic inputs used in the AI/ML development and validation processes may undermine trust in the safety of the cleared AI/ML enabled SaMD (6,25,26).

Furthermore, peer-reviewed publications were available for only 13/98 devices, among which only 2 were prospective studies evaluating a cleared CAD device. The importance of clinical evaluation, ideally through prospective studies, to externally test AI algorithms have been previously reported (27). Despite the FDA’s emphasis on prospective validation of cleared devices, developers are not incentivized to participate in head-to-head comparison of AI/ML devices . A recent study by Van Leeuwen etal, reported that only 9 of19 developers of AI/ML medical devices agreed to participate in a prospective multi-site evaluations in Europe(17). In this study, the other 11 studies were retrospective comparisons of their algorithmic output to earlier findings (28,29). Using prospectively collected data for clinical verification also offers greater transparency, which can further increase trust in both the device itself and its performance (15).

Finally, the mapped predicate lineages of all cleared CAD devices revealed that 6/8 CADx and 4/18 CADe/x devices showed mismatches in clinical function from their claimed predicate. Although the regulatory class created for de novo device DEN170022, from which all subsequent CADx devices were cleared, was broadly defined to cover all lesions suspected of cancer, the clinical function mismatches observed raises concerns about how truly equivalent a suspicious breast lesion is to an intracranial hemorrhage. Similarly, 12 breast lesion detection devices were cleared from the de novo DEN180005 for broadly detecting bone fractures. Such deviations can lead to questions about device safety – especially if no transparent image annotation instructions are provided in the 510(k) summary. Without access to such algorithmic input details, users are severely limited in their ability to objectively evaluate whether the claimed equivalency of the cleared device is valid.

This study highlights the challenges inherent in the current 510(k) pathway for ensuring safety and effectiveness of AI/ML-enabled radiological SaMD, particularly regarding transparency of the algorithm development process. The limitations in transparency of algorithmic input suggest that additional reporting mechanisms are needed ensure public trust in AI/ML-enabled devices. While it is evident that the FDA is adapting existing frameworks to the best of its purview, the current 510(k) pathway was originally developed for non-SAMD products. As such, it may not be optimally suited to accommodate the rapid innovation in AI/ML SaMD and stakeholders’ needs, where tensions balancing regulatory requirements and innovation is likely. One approach to address existing concerns about AI/ML radiological medical devices could involve greater transparency from developers. Specifically, developers need to provide the minimal yet key details that enables clinicians and evaluators to make informed decisions regarding the safe and effectiveness of FDA-cleared AI/ML SaMD.

may help ease safety concerns that limit adoption of AI. Even publicly available full predicate maps would allow the public to more easily assess the level of equivalence between each device and its claimed predicate.

There are a few limitations to this study. First, the FDA’s 510(k) database of AI/ML medical devices may not include recently-cleared devices. Second, the AI/ML devices were identified solely based on the descriptions within the 510(k) summaries and associated publications, which varied significantly in detail. This variability raises the possibility that some devices might have been misclassified as AI/ML medical devices based on how image labeling was performed. Third, despite a thorough search for related peer-reviewed publications on each product’s webpage, some relevant publications may have been overlooked.

## Conclusion

This study highlights a lack of transparency in key algorithm development details for radiological AI/ML-enabled SaMD cleared through the current FDA 510(k) process. The notable mismatches in clinical function between cleared devices and their claimed predicates may lead to public mistrust in device safety, as there is no publicly available data to validate the degree of true equivalence between devices. New mechanisms for increased transparency and more easily accessible information may help the public more easily assess the degree of claimed substantial equivalency, especially when prospective studies are rarely conducted. Such transparency could improve trust in AI/ML devices and may promote the adoption of radiological AI/ML-enabled devices in clinical settings.

## Data Availability

All data produced in the present work are contained in the manuscript

