## Appendix.A for "Beyond the Black Box: Avenues for Transparency in Regulating Radiological AI/ML-enabled SaMD via the FDA 510(k) Pathway"

**Appendix: Definition of variables used to index FDA Cleared AI/ML Radiological Devices**

| Variable | Definition |
| --- | --- |
| Entry Date | The date the observation was collected and put into our data set, not ACR Data Science Institute AI Central. |
| Date Cleared | The date the device was cleared by the FDA |
| Product Name | The name of product as listed in the "Trade/Device Name" field of the 510(k) Summary. Assigned in the "Product" column by the ACR Data Science Institute AI Central |
| Developer | The name of the company that filed the 510(k). |
| Type | Delineates the type of algorithm function into: Image Processing/Quantification, CADt, CADe, CADx and CADe/x. Assigned by the ACR Data Science Institute AI Central |
| Diagnosis | The general medical diagnosis which the device assists in producing. Assigned by the ACR Data Science Institute AI Central |
| Body Area | The area of the body to which the device is related. Assigned by the ACR Data Science Institute AI Central |
| Subspeciality | The general medical subspecialty to which the devices function applies. Assigned by the ACR Data Science Institute AI Central |
| Imaging Modality | The imaging method(s) the device utilizes. Assigned by the ACR Data Science Institute AI Central |
| Age Target Population | The age group for which the device is intended to treat. Assigned by the ACR Data Science Institute AI Central |
| Algorithmic Input: Image labeling instructions or description in FDA 510(k) document | There exists any reference to how the training data was created and labeled on either the 510(k) summary or the developer website. Essentially, if the training data is at all mentioned. |

| | |
|---|---|
| Algorithmic Input: Image labeling or description on product website or relevant publication | On the developer's website, there exists a set of instructions detailing how the training data was annotated. |
| Ground Truth | Quotation of the 510(k) summary document where the method of obtaining the ground truth specifically for the validation testing of the device is stated. The purpose of this variable is to capture the metric by which the performance of the algorithm is measured; therefore, ground truth for clinical validation (i.e. how a radiologist's performance changes as a result of using the device) is not counted for this field. |
| Developer Website information | Developer website contains a webpage that mentions the device by its name as it appears in the "Trade/Device Name" field of the 510(k) summary document or that is otherwise indicated to represent the device described in the 510(k) with either a link to the 510(k) itself, the 510(k) number, or a statement indicating that the device is FDA cleared which reasonably implies that the device is described by the 510(k) |
| Webpage Link | Product webpage exist |
| Publications on product webpage | There exists a link to a published, peer-reviewed article about the device on the webpage that most specifically presents the device within in the producer's website. |
